# Waning of SARS-CoV-2 Antibody levels response to inactivated cellular vaccine over 6 months among healthcare workers

**DOI:** 10.1101/2021.12.30.21268532

**Authors:** Monica Taminato, Ana Paula Cunha Chaves, Richarlisson Borges de Morais, Luiz Vinicius Leão Moreira, Danielle Dias Conte, Klinger Soares Faico Filho, Maria Cristina Gabrielloni, Eduardo Alexandrino de Medeiros, Nancy Bellei

**Affiliations:** Paulista Nursing School, Federal University of Sao Paulo, Sao Paulo, Brazil; Department of Medicine, Discipline of Infectious Diseases, Federal University of Sao Paulo, Sao Paulo, Brazil; Technical School of Health, Federal University of Uberlandia, Minas Gerais, Brazil; Infectious Diseases Division, Infection Control Committee, Hospital São Paulo, Federal University of Sao Paulo, Sao Paulo, Brazil

**Keywords:** COVID-19, Infectious disease, Infection prevention and control, Vaccine, Antibodies, Healthcare workers

## Abstract

**Background:** Health Care workers (HCW) are an important group affected by this pandemic and COVID-19 has presented substantial challenges for health professionals and health systems in many countries. The Brazilian vaccination plan implemented in October, so that third dose for HCW. However, the persistence of CoronaVac vaccine-induced immunity is unknown, and immunogenicity according to age cohorts may differ among individuals.

**Objective:** Evaluate the post vaccination immune humoral response and the relationship between post-vaccination seropositivity rates and demographic data among Healthcare Workers over 6 months after CoronaVac immunization.

**Methods:** A cross section study including Healthcare professionals vaccinated with CoronaVac for 6 months or more. The study was carried with the analysis of post-vaccination serological test to assess the levels of humoral response after vaccination.

**Results:** 329 participants were included. Among them, 76% were female. Overall, 18.5% were positive quantitative titles (IQR 42.87-125.5) and the negative group was 80%, quantitative titles (IQR 5.50-13.92).

**Conclusion:** It was possible to identify a group with positive quantitative titles in serological test for IgG antibody against the SARS-CoV-2. Further investigation is required to determine the durability of post-vaccination antibodies and how serological tests can be determine the ideal timing of vaccine booster doses.

## INTRODUCTION

Severe acute respiratory syndrome coronavirus 2 (SARS-CoV-2) has infected millions of people around the world. Brazil is among the countries with the highest numbers of confirmed cases and deaths from SARS-CoV-2. ^(1,2)^ Health care workers (HCWs) are an important group affected by this pandemic, and COVID-19 has presented substantial challenges for health professionals and health systems in many countries. They are also at high risk of infection with severe acute respiratory syndrome coronavirus 2 (SARS-CoV-2) due to direct contact with infected patients and the stresses of an overwhelming burden of labour required from them. ^(3)^

The CoronaVac (Sinovac Life Sciences, Beijing, China), an inactivated vaccine, was approved for emergency use by ANVISA, Brazil Ministry of Health on January 17, and was the first formulation distributed soon after authorization on January 18, 2021. ^(4)^ HCWs received two doses of the CoronaVac vaccine with the recommended dosing interval of 28 days between the first and second doses, the schedule considered to induce the highest effectiveness against the more severe outcomes of hospitalization, ICU admission, and death. ^(5)^

The decline in serum antibodies against SARS-CoV-2 observed in some studies raises questions about long-term immunity. Lower antibody levels are associated with new cases of Covid-19 even after vaccination, leading to the consideration of booster doses. ^(6-8)^ The SARS-CoV-2 seropositivity over time might be associated with the risk of future infection, since studies have shown that neutralizing and binding antibodies show a strong correlation with efficacy.^(6)^

The Brazilian vaccination plan was implemented in October, so that a third dose for HCWs and people aged 60 years and over was administered. However, the persistence of CoronaVac, vaccine-induced immunity is unknown, and immunogenicity according to age cohorts may differ among individuals. This study aimed to evaluate the post-vaccination immune humoral response, and the relationship between post-vaccination seropositivity rates and demographic data, (age and sex) among HCWs at > 6 months after their CoronaVac immunization.

## METHODS

We performed a cross section study including healthcare professionals from São Paulo Hospital, vaccinated with CoronaVac for 6 months or more. The study was carried out in October with an analysis of post-vaccination IgG antibodies.

### Subjects

São Paulo Hospital healthcare workers were invited to collect blood samples for serological tests to determine the quantitative anti-RDB IgG to assess the levels of humoral response after at least six months of vaccination by the whole-virion CoronaVac (Sinovac Life Sciences). All professionals who had received the second dose of the CoronaVac vaccine more than 6 months previously were eligible. Exclusion criteria were a previous infection with COVID-19, immunosuppression or use of immunosuppressive drugs. The study was approved by the Ethics Committee of the Federal University of São Paulo (CAE-47617621.6.0000.5505). All participants signed an informed consent form and participated in the study. A total of 334 HCW were garnered, aged 19-86 years.

### Laboratory study

The study was carried out in October 2021 in the Virology Laboratory. Briefly, three to five mL of venous blood was taken from the volunteers participating in the study. Sera were separated and stored in a -20°C freezer until the antibody studies were performed. IgG antibody assays against SARS-CoV-2 RBD protein were performed using the Access SARS-CoV-2 IgG antibody (1°IS) (Beckman Coulter, Inc.) according to the manufacturer’s instructions. The antibodies against the RBD of the spike protein were quantitatively analysed and were interpreted as positive (signal for test sample/signal at cut-off value if ⋝ 30UI/mL or BAU/mL) or negative (if ⋜ 30UI/mL) in accordance with the manufacturer’s instructions for the Access SARS-CoV-2 IgG Antibody Test.

### Statistical analysis

Data are presented as counts, percentages, and 95% confidence intervals (CIs). Comparisons were obtained between positive and negative groups according to the detection of IgG anti-RBD. The Shapiro-Wilk normality test was performed to verify normality, and the significance level was set at 5%. The Kruskal–Wallis test, also with a 5% significant level, was used for variables that did not follow normality. When there was significance, the Dunn test was used to check for multiple comparisons.

## RESULTS

Among the 329 participants, 251 (76%) were female and the median age was 41 years (19-85; IQR 31-52.5). Four patients included in the study tested negative, and were not included in the age cohort analysis. The seropositivity was 17.9% for females and 20.5% for male HCWs.

Overall, 18.5% (61) of the participants’ results were positive with a 64.47 BAU/mL anti –RDB IgG median quantitative titer (IQR 42.87-125.5) obtained for the whole study group. The minimum and maximum titers obtained for the positive samples were 30.16 – 1094 BAU/mL. Three participants presented with a titer above 506 BAU/mL, a titer previously considered as a correlate of 80% vaccine efficacy against symptomatic SARS-CoV-2 infections. The negative group included 80% (268) of the participants with a 8.55 anti –RDB IgG median quantitative titer (IQR 5.5-13.92) and the maximum titer was 29.92 BAU/mL (p <0.001).

IgG titers obtained for female HCWs were not different from those obtained for the male participants with a seropositivity test of 62.93; IQR 42.33-110.0 and 73.02 IQR 49.79 -154.0 BAU/mL respectively (p=0,296).

The testing volumes, number of positive results, and IgG titers in each age group are shown in the Table 1 and Figure 1 below.

**Table 1.** IgG anti-RBD seropositivity and antibody titers detection after 6 months of immunization with 2 doses of CoronaVac vaccine in HCW

| Age (years) | No.<sup>a</sup> | Positive – No. (%) | IgG titers med.<sup>b</sup> – BAU/ml<sup>c</sup> (IQR<sup>d</sup>) | OR<sup>e</sup> (95% CI) | P value<sup>*</sup> | ANOVA<sup>**</sup> |
| --- | --- | --- | --- | --- | --- | --- |
| **TOTAL** | 325 | 61 (18.26) | 64.47 (42.87-125.5) | - | - | - |
| **20-30** | 76 | 10 (13.15) | 74.06 (46.80-94.37) | Ref.<sup>f</sup> | Ref. | Ref. |
| **31-40** | 74 | 18 (24.32) | 62.85 (44.9-102.00) | 0.47 (0.20-1.10) | 0.09 | 0.99 |
| **41-50** | 72 | 13 (18.06) | 105.80 (47.00-503.30) | 0.68 (0.28-1.68) | 0.4 | 0.0145 |
| **>51** | 103 | 20 (19.42) | 51.73 (40.67-137.10) | 0.62 (0.27-1.43) | 0.32 | 0.99 |

<sup>a</sup> Number of cases
<sup>b</sup> Median
<sup>c</sup> Binding Antibodies Units per milliliter
<sup>d</sup> Interquartile range
<sup>e</sup> Odds ratio
<sup>f</sup> Reference
<sup>\*</sup> Fisher's exact test
<sup>\*\*</sup> Multiple comparison ANOVA/ Tukey's test and Dunn test.

**Figure 1.**
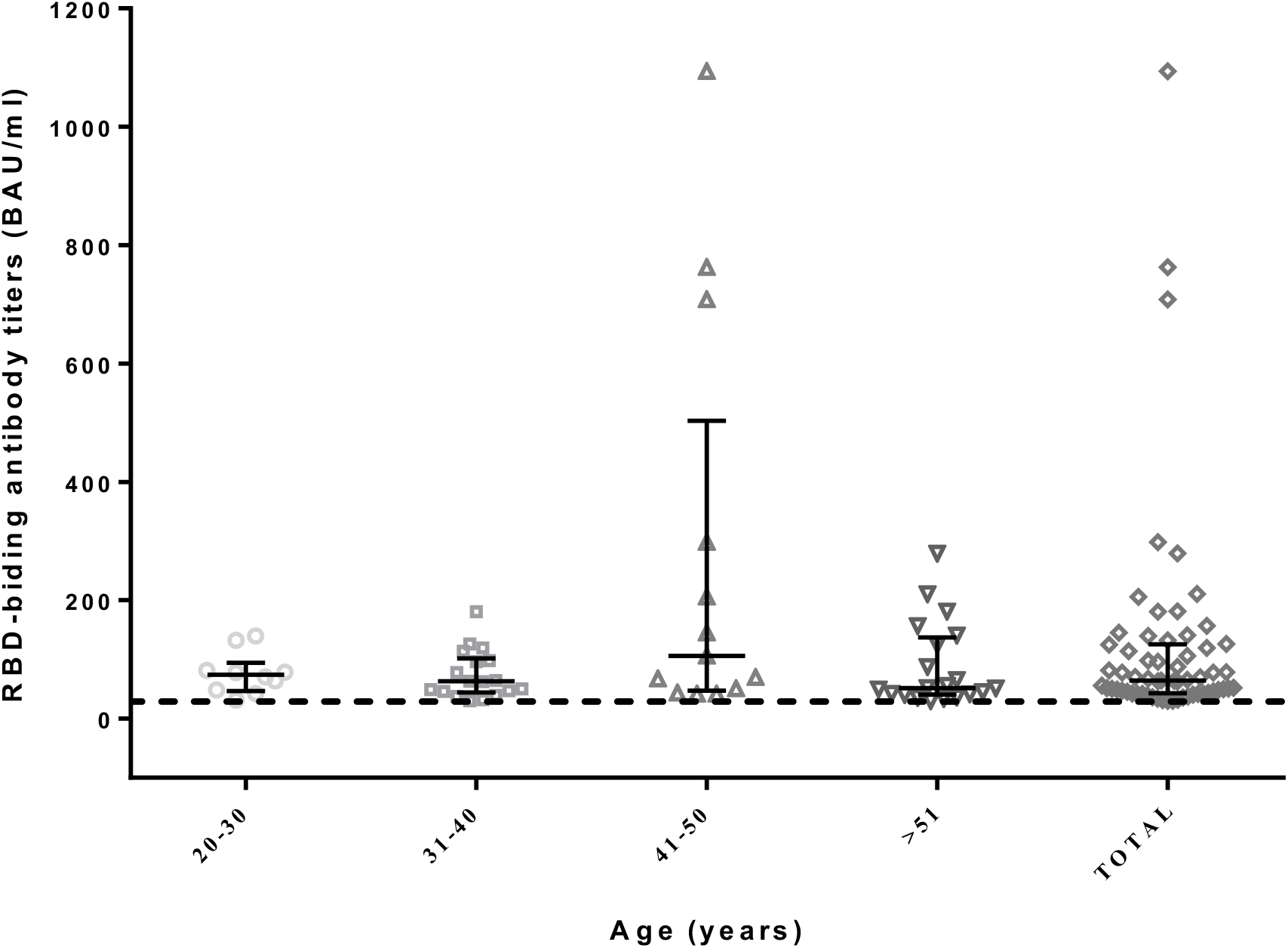
Comparision of Severe Acute Respiratory Virus 2 anti-RBD antibodies titers in HCW according to age groups. Dashed line indicates cut-off for seropositivity (≥ 30 BAU/ml). Vertical solid lines indicate the distance between the interquatile ranges. The middle horizontal solid lines indicate the median of antiboties titers, meanwhile the upper and lower horizontal edges representes the 25% and 75% percentiles of antibodies levels.

## DISCUSSION

The major challenge, at present, is to develop predictive models of immunological protection for COVID-19 and to define the correlates of protection to establish vaccination programs. ^(7,9)^

Data from previous studies have suggested that a 28 day dosage interval reaches a more robust antibody response, and a longer persistence, compared to a scheme with an interval of 14 days. ^(10)^ Our study found that only 20% of the evaluated healthcare professionals without previous SARS-CoV-2 infections vaccinated for more than 6 months with the two doses of the CoronaVac vaccine were seropositive.

Recent research has suggested that a high level of neutralizing titers are required to protect against severity and death from circulating SAR-CoV-2 variants. ^(11)^ We have observed a decrease in antibody detection without significant differences among different age groups. Indeed, there were no statistical differences between age and sex regarding IgG levels obtained more than 6 months after the CoronaVac vaccine. Only three participants obtained a high level of antibody titer that was considered to be correlated with protection, according to a recently published study. ^(9)^ The authors evaluated immunological markers 28 days after the second dose of ChAdOx1 vaccine and the risk of symptomatic COVID-19 decreased with increasing levels of anti-spike anti-RBD IgG. In their study, the antibody level needed in order to obtain an 80% efficacy of the VE vaccine against symptoms, was a mean of 506 (95% BAU / ml10.) Another report evaluated an immune correlate analysis of the mRNA-1273 COVID-19 vaccine trial and estimated a 90% vaccine efficacy of 57 RBD IgG level of 775 BAU/ml at the time. ^(12)^ In this view, even our seropositive HCW would not be protected against symptomatic infection at this moment.

One limitation of this study is the use of an immunochemiluminescence test as a surrogate marker of the immune humoral response, and not a plaque reduction neutralization test. Nonetheless, the assay detected the total immunodominant neutralizing antibodies that targeted the viral peak protein (S) receptor binding domain (RBD). It is generally used as a test of high sensitivity and specificity, and is evaluated as a correlate of protection in the recent studies cited above. Indeed, a previous study on IgG antibody tests and their correlation with the SARS-CoV-2 surrogate virus neutralization test (sVNT) in patients with COVID-19 has been evaluated and validated by analyzing convalescent serum and samples from non-COVID-19 patients. ^(13)^

Other immunological markers not tested in our study may contribute to the protection of previously immunized patients, even in the absence of antibody persistence. A non-peer-reviewed study of Brazilian professional health workers reported a 50.7% efficacy of CoronaVac in the prevention of severe forms of SARS-CoV-2 infections in a phase 3 clinical trial. ^(14)^ Another study published in China showed that HCW maintained their B cells and T cells specific for SARS-CoV-2 detection five months after two doses of the Sinopharm vaccine. ^(15)^

## CONCLUSION

Further investigation is required to determine the durability of post-vaccination antibodies in individuals, including other immunologic markers, and to elucidate how serological tests can be predictive of effectiveness and determine the ideal timing of vaccine booster doses for population protection.

## Data Availability

All data produced in the present study are available upon reasonable request to the authors.

